# Utilizing novel fluorothymidine PET imaging in a phase I study of veliparib on an intermittent and continuous schedule given in combination with carboplatin in metastatic breast cancer

**DOI:** 10.1101/2020.01.20.20018218

**Authors:** Robert Wesolowski, Daniel G. Stover, Maryam B. Lustberg, Abigail Shoben, Meng Zhao, Ewa Mrozek, Rachel M. Layman, Erin Macrae, Wenrui Duan, Jun Zhang, Nathan Hall, Chadwick L. Wright, Katharina Schregel, Susan Gillespie, Michael Berger, Andrea Camp, Jeffrey J. Chalmers, Priya Balasubramanian, Brandon L. Miller, Peter Amaya, Eleni Andreopoulou, Joseph Sparano, Charles L. Shapiro, Miguel Angel Villalona-Calero, Susan Geyer, Alice Chen, Michael R. Grever, Michael V Knopp, Bhuvaneswari Ramaswamy

## Abstract

Poly(ADP-ribose) polymerase inhibitors are FDA-approved for treatment of *BRCA* mutated metastatic breast cancer (MBC). Prior studies demonstrated benefit of adding oral PARPi veliparib to carboplatin and paclitaxel in *BRCA* mutation positive patients with MBC. We sought to find the recommended phase II dose (RP2D) and schedule of veliparib in combination with carboplatin in patients with advanced breast cancer, either triple negative (TNBC) or hormone receptor (HR) positive, HER2-negative with defective functional Fanconi Anemia (FA) DNA-repair pathway. Patients received escalating doses of veliparib on a 7, 14, or 21-day schedule with carboplatin every 3 weeks. Patients underwent [18]fluoro-3’-deoxythymidine (^18^FLT) positron emission tomography (PET) imaging, assessed in a blinded fashion. Forty-four patients (39 TNBC, 5 HR-positive/HER2-negative with a defective FA pathway) received a median of 5 cycles (range 1-36). Observed DLTs were grade (G) 4 thrombocytopenia (N=4), G4 neutropenia (N=1) and G3 akathisia (N=1). Common grade 3-4 toxicities included thrombocytopenia, lymphopenia, neutropenia, anemia, and fatigue. Of 43 patients evaluable for response, 18.6% achieved partial response and 48.8% had stable disease. Median progression free survival was 18.3 weeks. RP2D of veliparib was established at 250 mg twice daily on days 1-21 along with carboplatin at AUC 5. Patients with partial response had significant drop in SUV_max_ of target lesions between baseline and early ^18^FLT-PET (day 7-21; p_trend_=0.006). Continuous dosing of veliparib and every three week carboplatin demonstrated activity and acceptable toxicity. Decrease in SUV_max_ on ^18^FLT-PET scan during the first cycle of this therapy can identify patients likely to have a response.

**IMPLICATIONS FOR PRACTICE:** The BROCADE studies suggest that patients with *BRCA* mutation benefit from addition of PARP inhibitor veliparib to carboplatin plus paclitaxel. In this study, we demonstrate that a higher dose of veliparib is tolerable and active in combination with carboplatin alone. With growing interest in imaging-based early response assessment, we demonstrate that decrease in ^18^FLT-PET SUV_max_ in the first cycle of therapy is significantly associated with response. Collectively, this study provides clarity on dosing of veliparib with carboplatin in advanced breast cancer while providing additional data on the potential for novel PET imaging modalities in monitoring therapy response.

## Introduction

Poly (ADP-Ribose) Polymerase (PARP) protein family consist of 17 enzymes that have several key functions during DNA repair. PARP proteins sense single strand DNA breaks (SSB), signal the presence of DNA damage, generate linear and branched poly(ADP-ribose) chains, recognize topoisomerase I cleavage complexes and facilitate base excision repair (BER).^1-3^ PARP-1 and PARP-2 are considered as the primary enzymes involved in the repair of single-stranded DNA breaks through the BER pathways.^4^ Poly(ADP-ribosyl)ation (PAR) has been implicated in many cellular processes and enhanced PARP-1 expression and/or activity is one of the mechanisms by which tumor cells evade apoptosis caused by DNA damaging agents.^5,6^

BRCA1 and BRCA2 proteins are essential for homologous recombination repair. BRCA dysfunction renders cancer cells dependent on BER and hence increased sensitivity to PARP inhibition (PARPi).^7,8^ In pre-clinical studies, BRCA deficient cells are also more sensitive to platinum drugs than BRCA proficient counterparts both *in vitro* and *in vivo* and combining PARPi and platinum agents was shown to be synergistic providing a rationale to study these combinations in clinic.^9,10^ Clinical trials studying regimens containing platinum regimens have also demonstrated that patients with triple negative breast cancer respond well to these agents.^11-13^ Therefore, this provides a rationale to study combinations of platinum drugs with PARP inhibitors in patients with advanced triple negative breast cancer and/or DNA repair defects.^14^

BRCA1/2 are part of the Fanconi Anemia (FA) network of proteins that function in DNA-damage response to maintain genome integrity, and include BRCA2/FANCD1, ataxia telangiectasia, Rad3 related protein (ATR), and BRCA1, among others^7,8,15,16,17^. The common hallmark of defective FA core complex such as FANCF methylation, is lack of ubiquitination of FANCD2, leading to lack of FANCD2 foci in the nuclei of the tumor cells in S phase.^18^ We hypothesized that breast tumors with defective DNA repair due to inactivation of BRCA/FA pathway, so-called “BRCAness” will be susceptible to treatment with platinum in combination with PARP inhibitor.^19^

3’-deoxy-3’-[F-18] fluorothymidine ([F-18]FLT) is a radiolabeled imaging agent with structural analog of the DNA constituent, thymidine.^20^ The activity of this radiolabeling agent is dependent on cells undergoing DNA replication and hence ^18^FLT uptake is dependent on the proliferative rate of the cells. This is in contrast to a more conventional 5-fluorodeoxyglucose (^18^FDG) PET scan where uptake of FDG depends on high intake of glucose reflective of increased metabolic rate. There is growing interest in PET-based imaging for early response monitoring in breast cancer^21,22^ and uptake of ^18^FLT during therapy with PARP inhibitors and DNA damaging agents lends itself well as an attractive imaging modality to study changes in DNA synthesis and early response to this therapy.

This multicenter phase I study (NCI8609) sponsored by Cancer Therapy Evaluation Program (CTEP) at the National Cancer Institute (NCI), sought to assess the recommended phase 2 dose (RP2D) of veliparib on an intermittent (7 or 14 day) or continuous (21 day) schedule in combination with every three week schedule of carboplatin in patients with advanced breast cancer that was either triple negative or HR positive (estrogen and/or progesterone receptor positive), HER2 negative with defective FA pathway based on lack of FANCD2 foci in the nuclei of proliferating tumor cells detected by FA Triple Stain Immunofluorescence (FATSI) assay. We report the primary endpoint of RP2D and schedule of veliparib in combination with carboplatin, secondary endpoints of efficacy, and exploratory endpoints of ^18^FLT-PET scans.

## Methods

### Patients

Patients eligible for the trial were adult women with metastatic or locally advanced inoperable breast cancer which fulfilled one of the three criteria: 1) negative for estrogen (ER), progesterone (PR) and HER2 receptors (based on ASCO/CAP guidelines); 2) HR positive (defined as ER and/or PR positive), HER2 negative breast cancer that is deficient for the FA pathway based on the FA Triple Stain Immunofluorescence (FATSI) test (i.e. no FANCD2 foci in nuclei of 100 proliferating tumor cells); 3) HER2 negative breast cancer with known germline *BRCA1/2* mutation. HR positive/HER2 negative patients without known germline BRCA 1 or 2 mutation initially signed a screening consent for testing their archival tumors for FATSI and only proceeded with the therapeutic portion of the study if testing showed deficiency in FA pathway. Other eligibility criteria requirements included no more than three prior chemotherapy regimens for metastatic disease and Eastern Cooperative Oncology Group (ECOG) performance status of 0 to 2. Patients with treated CNS metastasis were eligible. Prior platinum exposure was allowed.

### Ethics

All patients provided written informed consent. This study was approved by the local institutional review boards at Ohio State University and each participating site and conducted in accordance with Good Clinical Practice guidelines and the Declaration of Helsinki. This trial was registered with ClinicalTrials.gov on December 2, 2010 with identifier NCT01251874.

### Study Design and Treatment

This was a multicenter, CTEP-sponsored, single-arm phase I trial of veliparib on an intermittent (7 or 14 day) or continuous (21-day) schedule given in combination with carboplatin in patients with advanced breast cancer. The study used a standard 3+3 dose escalation design. The primary objective was to determine recommended phase II dose of veliparib in combination with carboplatin defined as the maximum tolerated dose (MTD) or the highest dose level (if MTD could not be determined). Other objectives included assessment of safety and tolerability and preliminary efficacy of the combination. Veliparib was initiated at 50 mg, twice daily (BID), orally for 1-7 days of 21-day cycles (dose level 1 and 1A). If tolerated, the schedule of veliparib was escalated to days 1-14 of a 21-day cycle (dose levels 2-5) and then to continuous dosing (dose levels 6-7). Dose of carboplatin was held stable in all dose levels at area under the curve (AUC) of 5 mg/mL x minute (except for dose level I where the dose was AUC 6). Dose escalation of veliparib proceeded using a standard phase I dose escalation in cohorts of 3-6 patients for DL 1-7.

### Clinical Assessments

Dose-limiting toxicity (DLT) was defined as a significant adverse event occurring in the first cycle, and fulfilling one of the following criteria: grade ≥ 3 non-hematologic toxicity, electrolyte abnormalities of ≥ grade 3, grade 4 thrombocytopenia, febrile neutropenia, grade 4 neutropenia lasting for 7 days or more, or grade five toxicity. The MTD and the recommended phase 2 dose (RP2D) was defined as the highest dose at which no more than 1 out of six patients experienced a DLT. Adverse events (AE) were graded according to National Cancer Institute (NCI) Common Toxicity Criteria (version 3.0). Treatment could be delayed for up to 3 weeks to allow resolution of toxicities and a patient could have up to two dose reductions of veliparib, carboplatin, alone or concurrent. Tumor response were based on radiologist assessment according to the modified Response Evaluation Criteria in Solid Tumors (RECIST), version 1.1. A dedicated and blinded radiologist performed tumor assessments.

### Tumor tissue screening: Fanconi Anemia Triple Immunofluorescence Assay (FATSI)

Eligible patients with HR positive, HER2 negative breast cancer were consented to have their formalin fixed, paraffin embedded (FFPE) tumor tissue screened for FA functional deficiency using the FATSI test.^23^ The FATSI test employs a triple stain with Ki-67, DAPI and FANCD2 to identify FANCD2 foci in the nuclei of proliferating neoplastic cells. The assay was performed in a CLIA-certified laboratory.^23^ Negative FATSI test (i.e. absence of FANCD2 in the nucleus of 100 proliferating cells) would identify patients whose tumors were deficient in FA pathway.^24^

### Gamma (γ) H2 Ax assay

Circulating tumor cells (CTCs) were collected at baseline (day 1 and 3 of cycle 1) and serially (day 1, 7 and 14 of cycle 2), on day 1 of every 3 cycles and at progression, using negative selection technology based on immunomagnetic tagging and removal of CD45+ cells.^25,26^ We measured formation of γH2Ax in the CTCs using an antibody targeting the phospho-histone (Clone JBW301, Milipore Cat # 05-636).^27^ This primary mouse antibody was subsequently counter stained with a goat, anti-mouse IgG conjugated to Alexa Fluor 594 (Invitrogen, A-11005).

### FLT-PET to assess DNA damage induced by varying dose schedules of PARP inhibitor

We performed FLT-PET scan at baseline, cycle 1 day 7, day 14 (for cohorts receiving veliparib on days 1-14 of every cycle) or 21 (for cohorts treated with veliparib on days 1-21 of every cycle) and after cycle 3, day 1 to assess change in the uptake of FLT between patients with and without response. Lesions were track-matched with the FDG PET/CT and semi-quantitatively assessed using 2D ROI placement in a matched, blinded fashion.

### Statistical Methods

The safety population included patients who received at least one dose of study drug. Adverse events were summarized descriptively by attribution of study therapy (unlikely, probably, likely, and definitely related) and grade using Common Terminology Criteria for Adverse Events (CTCAE) version 3.0. Laboratory variables were summarized using mean change in value from baseline to scheduled time points for each dose level group and 95% confidence interval. Laboratory values were also categorized according to their CTCAE version 3.0 toxicity grade and tabulated by worst on-study toxicity grade and dose level group. Progression free survival was estimated using Kaplan Meier methods. All analyses conducted were using Stata for Windows and R version 3.4.1.

## RESULTS

### Patient demographics

Between December, 2010 and April, 2013, forty-four patients with metastatic or locally advanced inoperable breast cancer were enrolled from The Ohio State University Comprehensive Cancer Center and Montefiore Medical Center and received a median 5 cycles (range 1-36). Patients received upto three lines of prior chemotherapy regimens for metastatic disease. The baseline characteristics of the patients are outlined in **Table 1**. Thirty-nine patients had TNBC and five patients had HR positive/HER2 negative metastatic breast cancer with functional deficiency of FA pathway based on negative FATSI assay. Thirty-four TNBC tumors were tested for FA deficiency using the FATSI test. Of these four (11.8%) were found to have functional deficiency of FA pathway.

**Table 1.** Patient Demographics.

| Characteristics | Number of patients (%) |
| --- | --- |
|  | N= 44 |
| Median age in years (range) | 58 (31 – 77) |
| ECOG PS |  |
| 0-1 | 39 (89) |
| 2 | 5 (11) |
| ER/PR status |  |
| ER/PR - | 39 (89) |
| ER+ and/or PR+ | 5 (11) |
| # metastatic sites |  |
| 1-3 | 38 (86) |
| > 3 | 6 (14) |
| # prior chemo regimens (metastatic) |  |
| 0-1 | 29(66) |
| 2 | 7 (16) |
| 3 | 8 (18) |
| Prior platinum exposure | 8 (18) |
| BRCA 1/2 mutation | 7 (16) |
| Defect FA pathway | 9 (20) |

### Dose-limiting toxicities and Safety

All patients were evaluable for toxicity from the time of their first treatment with veliparib. Three patients were enrolled on dose level 1 (Veliparib 50 mg BID for 7 days) with carboplatin at area under the curve (AUC) 6 every 21 days. One patient developed grade (G) 4 thrombocytopenia and the dose level expanded to six patients. Two more patients in this dose level developed DLT (G4 thrombocytopenia and G4 neutropenia). The protocol was subsequently amended and the carboplatin dose was reduced to AUC 5 for all subsequent dose levels. No DLTs were observed in the three patients subsequently enrolled to dose level 1A with veliparib 50 mg BID for 7 days and carboplatin AUC 5. Patients were then enrolled on escalating doses of veliparib for 14 days and MTD was not reached at dose level 5 (**Table 2**). After discussions with the National Cancer Institute (NCI), veliparib schedule was changed to continuous dosing and two dose levels were planned with this schedule. No DLTs were observed at the highest planned dose of veliparib (250 mg BID for 21 days) in combination with carboplatin on day 1 (dose level 7). Dose escalation, number of patients in each cohort, and DLTs are outlined in **Table 2**. We did not observe any DLT on dose level 7, but four out of the six patients eventually required dose reductions of carboplatin and/or veliparib for thrombocytopenia or nausea. Therefore, the RP2D of veliparib in combination with carboplatin (AUC5 Q 3 weeks) was determined to be 250mg bid on a continuous schedule in dose level 7 (the highest dose level).

**Table 2.** Summary of dose levels and dose limiting toxicities.

| **Dose Level** | **Carboplatin Dose (AUC)** | **Veliparib Dose/Schedule (Days in each 21-day cycle)** | **N** | **Dose Limiting Toxicities** |
| --- | --- | --- | --- | --- |
| 1 | 6 | 50 mg BID (1-7) | 7 | G4 Thrombocytopenia (N=2)<br>G4 Neutropenia (N=1) |
| 1A | 5 | 50mg BID (1-7) | 3 | None |
| 2 | 5 | 50 mg BID (Days 1-14) | 6 | Gr 4 thrombocytopenia (N=1) |
| 3 | 5 | 100 mg BID (1-14) | 3 | None |
| 4 | 5 | 150 mg BID (1-14) | 6 | G3 Akathisia (N=1) |
| 5 | 5 | 200 mg BID (1-14) | 6 | G4 thrombocytopenia (N=1) |
| 6 | 5 | 200 mg BID (1-21) | 7 | G4 thrombocytopenia (N=1) |
| 7 | 5 | 250 mg BID (1-21) | 6 | None |

Fifty percent (N=22) of patients required dose reductions of either one or both agents primarily for myelosuppression (in particular thrombocytopenia). Thirty-three (75%) patients experienced at least one or more grade 3 or 4 toxicities, which were attributable to study treatment (**Table 3**). The most common and clinically significant grade 3-4 toxicity events were hematologic and included thrombocytopenia, neutropenia and anemia. Among non-hematologic toxicities that were G3 or higher, the most common were fatigue and vomiting (**Table 3**). No grade 5 toxicities were reported. Reasons for discontinuation of study therapy was disease progression (N=40) followed by patient withdrawal (N=1), adverse events (prolonged neutropenia) (N=1), and death due to disease progression on study (N=1).

**Table 3.**
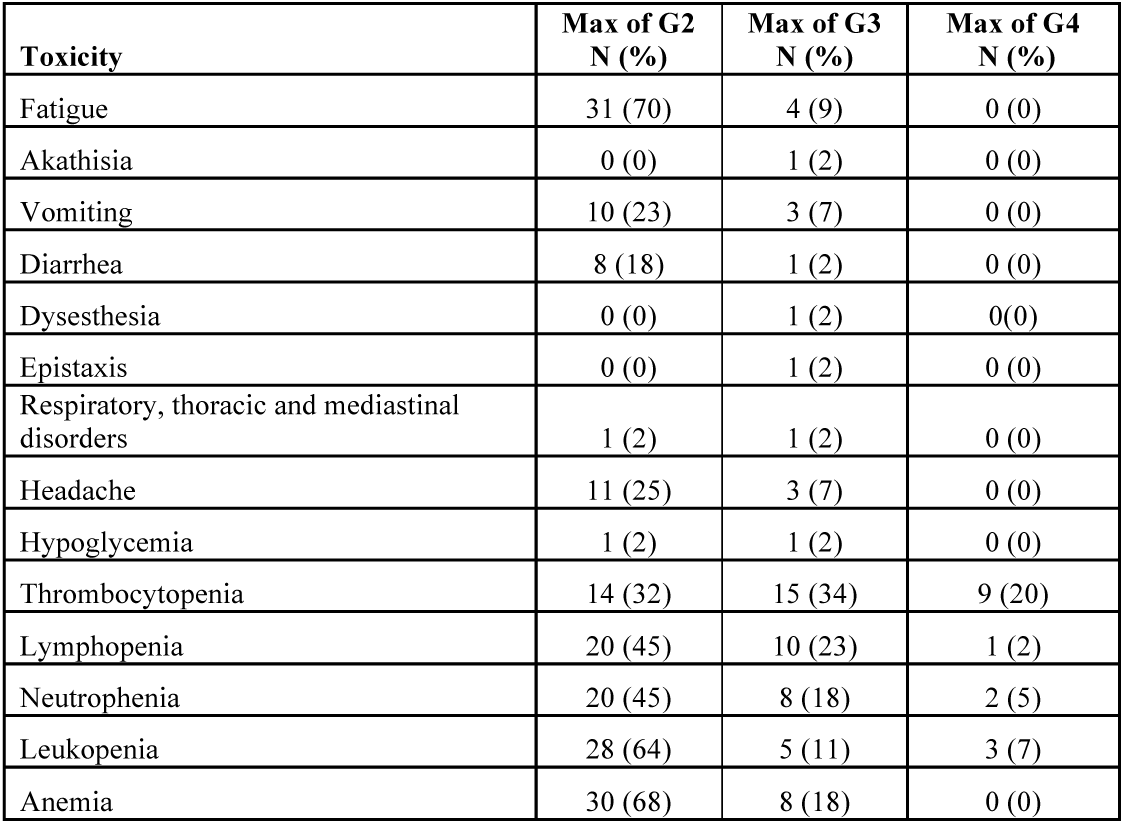
Overall number of patients with toxicity grade 2 or above.

| **Toxicity** | **Max of G2**<br>**N (%)** | **Max of G3**<br>**N (%)** | **Max of G4**<br>**N (%)** |
| --- | --- | --- | --- |
| Fatigue | 31 (70) | 4 (9) | 0 (0) |
| Akathisia | 0 (0) | 1 (2) | 0 (0) |
| Vomiting | 10 (23) | 3 (7) | 0 (0) |
| Diarrhea | 8 (18) | 1 (2) | 0 (0) |
| Dysesthesia | 0 (0) | 1 (2) | 0(0) |
| Epistaxis | 0 (0) | 1 (2) | 0 (0) |
| Respiratory, thoracic and mediastinal disorders | 1 (2) | 1 (2) | 0 (0) |
| Headache | 11 (25) | 3 (7) | 0 (0) |
| Hypoglycemia | 1 (2) | 1 (2) | 0 (0) |
| Thrombocytopenia | 14 (32) | 15 (34) | 9 (20) |
| Lymphopenia | 20 (45) | 10 (23) | 1 (2) |
| Neutrophenia | 20 (45) | 8 (18) | 2 (5) |
| Leukopenia | 28 (64) | 5 (11) | 3 (7) |
| Anemia | 30 (68) | 8 (18) | 0 (0) |

### Efficacy

Of 44 patients, one patient on DL 4 withdrew from the study after receiving only 2 cycles and was therefore not evaluable for response. Of the remaining 43 evaluable patients, 18.6% had a partial response (PR) (N=8); 48.8% had stable disease (SD) (N=21) as best response (**Figure 1A**). Of 21 patients with SD, 10 (23.3%) patients had SD > 24 weeks providing a clinical benefit rate (CBR) of 41.9%. The median progression free survival (PFS) for all patients who received at least 1 cycle of therapy was 18.3 weeks (95% CI: 10.9 to 22.0 weeks) and there was no significant difference in PFS across veliparib dosing schedule (log-rank p=0.87; **Figure 1B)**. Among the 8 patients with PR, median duration of response was 28.3 weeks (95% CI: 15.4 to 60.1 weeks). Median overall survival (OS) was 62.6 weeks (95% CI: 33.9 to 87.1 weeks; **Figure 1C**).

**Figure 1.**
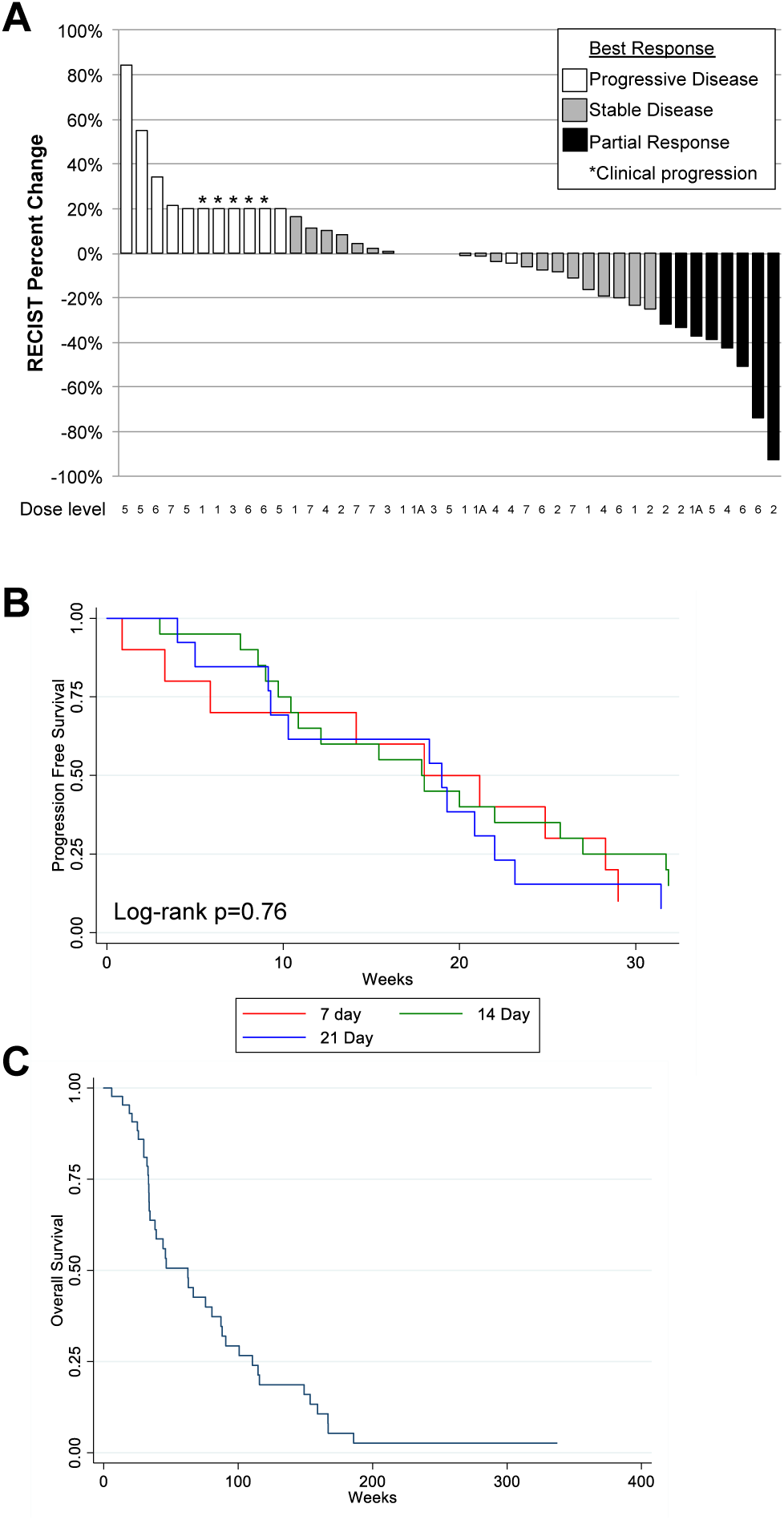
Efficacy and Outcomes. **A**. Waterfall plot of best response with RECIST percent change indicated on y-axis. Patients with clinical progression prior to cycle 3 are included at 20% fold change and indicated by asterisk (*). Best response indicated by color of bars: progressive disease, white; stable disease, grey; partial response, black. **B**. Progression-free survival from study entry by veliparib dosing schedule. Median progression free survival for all patients who received at least 1 cycle of therapy was 18.3 weeks. Veliparib dosing indicated by line color: 7-day dosing, red; 14-day dosing, green; 21-day dosing, blue. **C**. Overall survival from study entry. Median overall survival was 62.6 weeks.

### Deficiency in homologous recombination DNA repair and response

Of the nine patients with FATSI demonstrating defective FA pathway, 22.2% of patients achieved PR (N=2), 55.6% had SD (N=5) and 22.2% of patients (N=2) had primary progression. Four of the five patients with stable disease showed disease stabilization for > 24 weeks (44.4%). Among the 7 patients with known *BRCA1/2* mutation, 28.6% of patients (N=2) had PR, 71.4% of patients (N=5) had SD, and 42.9% of patients (N=3) SD > 6 months. When patients with tumors deficient in FA pathway based on FATSI testing and *BRCA1/2* mutations were analyzed together (N=16), 25% had a PR (N=4) and 62.5 % had stable disease (SD> 6 months occurred in 43.8% of patients). One patient with *BRCA1* mutation achieved durable partial response to study therapy and received total of 95 cycles of treatment. The patient was taken off study after developing thrombocytopenia and was subsequently diagnosed with myelodysplastic syndrome, assessed as possibly related to study therapy.

### ^18^FLT-PET Imaging: Correlation with ^18^FDG-PET and RECIST measurement

^18^FLT-PET imaging was obtained successfully in all patients treated at OSU with the expected proliferative whole body mapping revealing uptake in the bone marrow, liver and reticuloendothelial system (**Supplementary Figure 1**). There were no toxicities attributable to administration of ^18^FLT. The use of two distinct PET radiotracers in this study facilitates evaluation of distinct biological processes in cancer cells, specifically metabolic activity (^18^FDG-PET) and proliferation (^18^FLT-PET). We first evaluated the correlation between the two tracers (**Figure 2A**). Evaluating the primary target lesion at both baseline (BL) and first planned imaging (cycle 3 day 1; C3), we show that there is overall good correlation between SUV_max_ of ^18^FDG- and ^18^FLT-PET (Spearmon rho = 0.62, p=4.1e-07). However, the correlation <0.7 suggests that these tracers are not indistinguishable. We evaluated BL and C3 time-points independently and demonstrated similar correlations (**Supplementary Figure 2A/B**).

**Figure 2.**
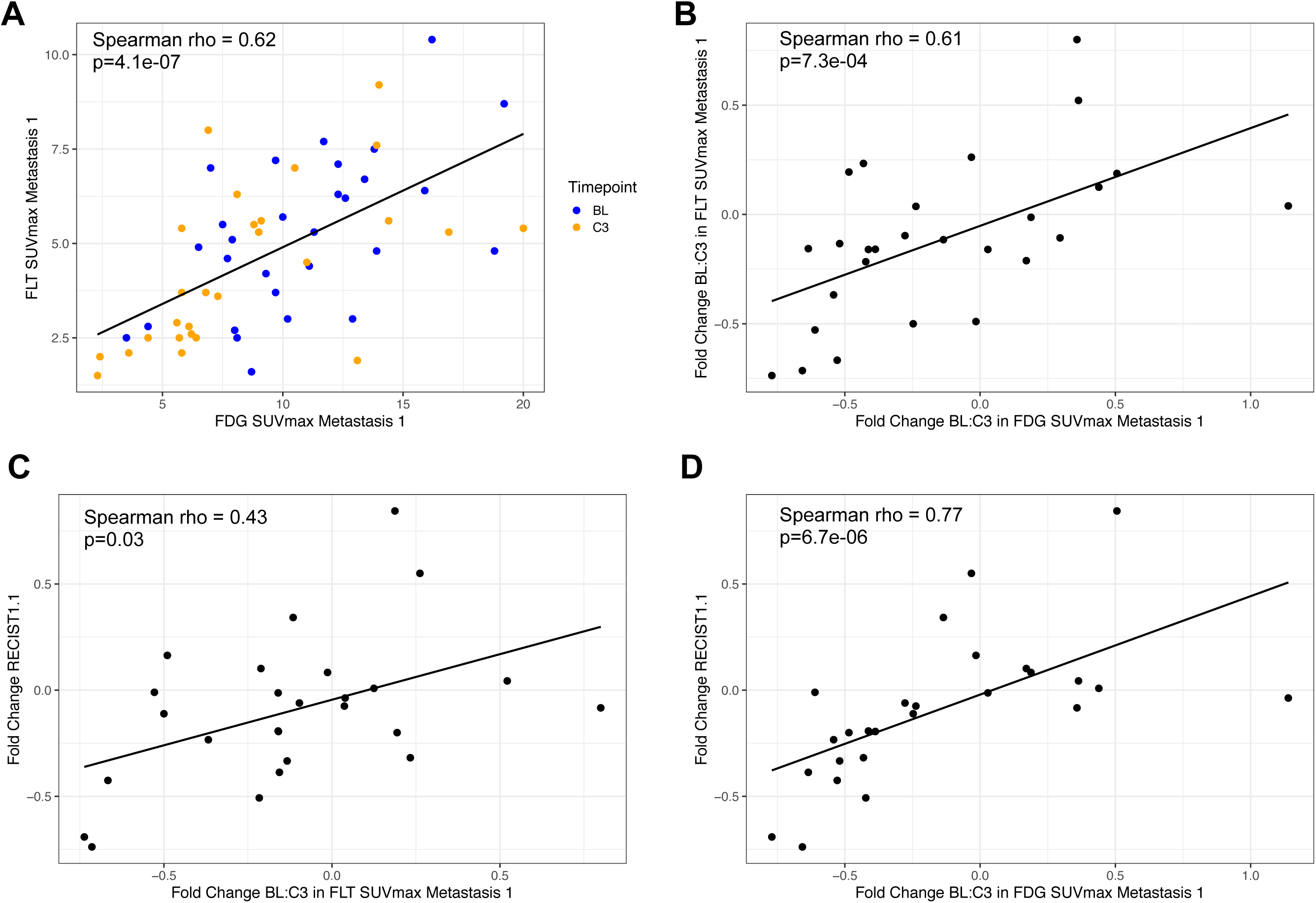
^18^FLT-PET Imaging: Correlation with ^18^FDG-PET and RECIST measurement. **(A)** Scatter plot of ^18^FLT-PET SUV_max_ of target lesion versus ^18^FDG-PET SUV_max_ of the same target lesion at baseline (“BL” ; indicated in blue) and cycle 3 day 1 (“C3” ; indicated in orange). Line of best fit indicated. **(B-D)** Scatter plot of fold change from BL to C3 of ^18^FLT-PET SUV_max_ of target lesion versus ^18^FDG-PET SUV_max_ of the same target lesion (**B)**, ^18^FLT-PET SUV_max_ of target lesion versus RECISTv1.1 (**C)**, and ^18^FDG-PET SUV_max_ of target lesion versus RECISTv1.1 measurement (**D)**. For all comparisons, correlation evaluated by Spearman’s rank correlation coefficient with p-value indicated.

To investigate dynamic changes over time on therapy, we evaluated the association of fold change from BL to first planned imaging (C3) for ^18^FLT-PET, ^18^FDG-PET, and RECISTv1.1 (**Figure 2B-D**). ^18^FLT-PET and ^18^FDG-PET primary target lesion SUV_max_ fold change from BL to C3 showed similar correlation to the simple SUV_max_ values (Spearmon rho = 0.61, p=7.3e-04). When comparing RECISTv1.1 measurements BL:C3 fold change to the PET metrics, ^18^FLT-PET primary target lesion SUV_max_ fold change showed lower correlation (Spearmon rho = 0.43, p=0.03) than ^18^FDG-PET primary target lesion SUV_max_ (Spearmon rho = 0.77, p=6.7e-06).

### Serial ^18^FLT-PET imaging and association with response

We performed ^18^FLT-PET scan at four time points: 1) BL; 2) cycle 1 day 7 (time-point 1; T1); 3) day 14 for cohorts receiving veliparib on days 1-14 of every cycle or day 21 for cohorts treated with veliparib on days 1-21 of every cycle (time-point 2; T2); and 4) after cycle 3 day 1 (C3) to assess change in the uptake of ^18^FLT between patients with and without response. The change in SUV_max_ on ^18^FLT-PET between baseline and follow up scans did not depend on dose or schedule of veliparib (N=24). Comparing the linear trend across 4 time-points in responders versus others, there was a statistically significant drop in ^18^FLT uptake in the responders (p=0.006) (**Figure 3A, top panel**). Among responders, patients had a rapid decrease in ^18^FLT uptake by T1 (cycle 1 day 7) with little change to T2 (cycle 1 day 14 or day 21) or C3 (**Figure 3A, bottom panel)**. As an exploratory analysis, we also evaluated fold change in ^18^FLT (**Supplementary Figure 2C/D)**. For responders (PR) versus non-responders (SD+PD), fold change from baseline to T1, T2, or C3 were all associated with response (nominal p<0.05) but not after multiple test correction (all FDR p>0.05), while fold change from T1 or T2 showed no association (**Supplementary Figure 2C**). We then evaluated ^18^FLT-PET SUV_max_ at BL versus C3, compared with ^18^FDG-PET SUV_max_ and RECISTv1.1 total measurement (**Figure 3B-D)**. At C3, all three metrics demonstrated significant drop among responders while those with stable disease or progressive disease did not show a significant decrease by any metric.

**Figure 3.**
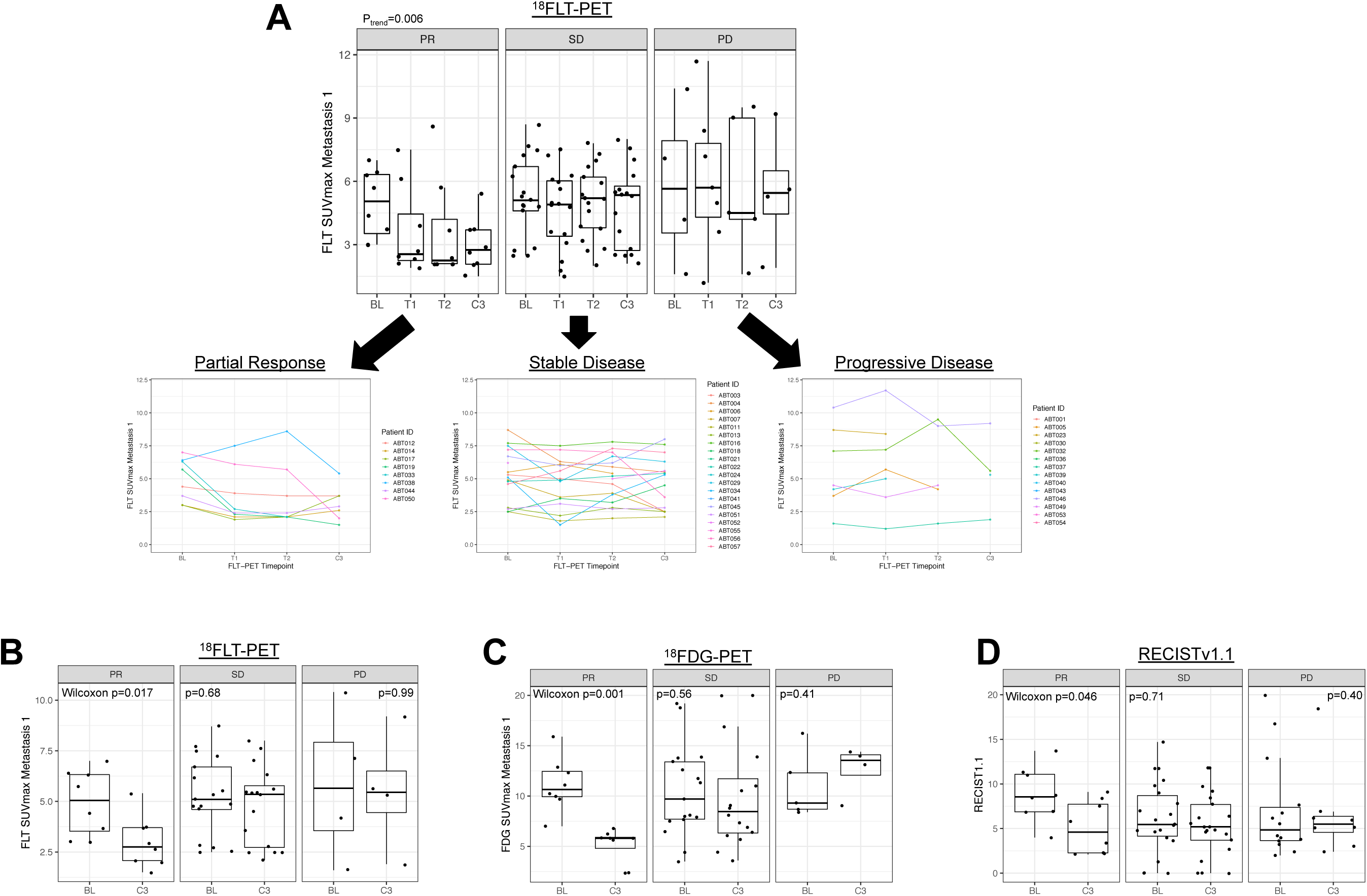
Early Change in ^18^FLT-PET SUV_max_. (**A**) **Top panel** shows ^18^FLT-PET SUV_max_ of target lesion from baseline (“BL”), to day 7 (“T1”) to day 14 or 21 timepoint (“T2”), to cycle 3 day 1 timepoint (“C3”). Patients are grouped by best response, partial response (PR), stable disease (SD), progressive disease (PD). The linear trend across 4 time-points in responders versus others (P_trend_) indicates was a trend towards the responders having an early drop in ^18^FLT uptake. **Bottom panel** shows individual patient level change at the four time points. **(B-D)** Baseline (“BL”) versus cycle 3 day 1 (“C3”) ^18^FLT-PET SUV_max_ of target lesion **(B)**, ^18^FDG-PET **(C)**, and RECISTv1.1 **(D)** grouped by response. P-value indicates Wilcoxon signed-rank test for each patient’s paired BL and C3 sample, by response grouping.

### Circulating tumor cells

Peripheral blood for CTCs were obtained serially in 36 patients enrolled at OSU with 32 patients having at least 3 serial samples. Although CTCs values did not have any correlation with response groups, gamma H2Ax in CTCs at baseline showed higher trend among those with a PR (p=0.02) and these values tended to be numerically higher during cycle 2 in this group (p=0.08), suggesting higher induction of DNA damage. (**Supplementary Figure 3**)

## DISCUSSION

Preclinical studies have shown that PARP1 inhibitors potentiate cytotoxicity when combined with platinum chemotherapy agents (cisplatin or carboplatin), which induce DNA damage through adducts and cross-linking.^28^ Veliparib (ABT-888) is an efficient oral PARP inhibitor that targets PARP1 and PARP2, the primary enzymes involved in DNA repair.^29-31^ Single agent PARP inhibitor was approved for patients with BRCA deficient hereditary advanced ovarian cancers and breast cancers.^32,33^ To date, no combinations of PARP inhibitors with other agents are FDA approved for breast cancer. Preclinical *BRCA* mutant models demonstrate the combination of veliparib and carboplatin as more effective than either drugs alone or combination of cisplatin plus veliparib^34^. The I-SPY 2 Trial showed that veliparib-carboplatin added to standard therapy resulted in higher rates of pathological complete response than standard therapy alone in TNBC.^11,35,36^ BrighTNess trial tested addition of veliparib to carboplatin or carboplatin alone to neo-adjuvant chemotherapy in patients with stage 2-3 triple negative breast cancer (not selected for BRCA 1/2 mutation) and showed that addition of carboplatin but not veliparib resulted in higher pathologic complete response rate.^35^ However, both these studies used low dose of veliparib at 50 mg PO twice daily on continuous schedule along with carboplatin in the early breast cancer setting.

Our multi-institutional phase 1 study demonstrated that veliparib in combination with carboplatin was well tolerated. Veliparib at 250 mg twice daily on a continuous schedule given along with carboplatin at AUC of 5 on day 1 of a 21-day cycle was the RP2D. Patients tolerated the combination well overall and the most common grade 3 or higher toxicities were hematologic. Our RP2D was higher than that was established by the California Consortium Trial (NCT01149083) in patients with *BRCA* mutation associated MBC, where the RP2D of veliparib with carboplatin was established at 150 mg BID (continuous dosing). The DLTs and toxicities reported in this study were similar.^37^ Another phase I study of veliparib combined with cisplatin and vinorelbine in advanced TNBC and/or BRCA mutation associated cancer established 300 mg BID (days 1-14 on a 21 day cycle) as the RP2D.^38^ Our trial is unique in not only focusing on all TNBC but also including HR positive patients with defective FA pathway. In addition, our phase I study investigated, three different schedules including, days 1-7, days 1-14 and continuous treatment of veliparib. Based on our study, in the metastatic setting, patients are able to tolerate higher dose of veliparib (250 mg daily) on a continuous schedule with carboplatin every three weeks provided that the dose of carboplatin is kept at AUC of 5.

The overall clinical benefit rate (CBR) in our patient population was 41.9% (CBR = PR + SD ≥ 6 months). Higher responses were seen in patients with germline *BRCA1/2* mutation (CBR 71.5%) and defective FA pathway (CBR 66.6%). This is similar to the response rates reported by other investigators. Somlo, et al reported a 51% CBR in their phase I study with veliparib and carboplatin in BRCA mutation carries with MBC.^37^ The phase II portion of this trial tested the efficacy of single agent veliparib at 300 mg BID in germline *BRCA1/2* mutation carriers, with those progressing on single agent veliparib treated with the combination carboplatin and veliparib at 150 mg BID. Interestingly, the median PFS of the 30 patients treated with this combination, post-progression on single agent veliparib was low at 1.8 moths (95% CI: 1.4 - 2.3). This suggests that combining veliparib with platinum agent earlier in disease course may be a better strategy. Another study, reported a 35% overall response rate in all patients (TNBC) but showed a 57% response rate amongst patients with BRCA mutation treated with the combination of cisplatin, vinorelbine and veliparib.^38^ Phase II trial of combination of paclitaxel, carboplatin and veliparib in BRCA2 carriers resulted in a 77.8% response rate (BROCADE).^39^ Subsequent phase III, double blind randomized study (BROCADE3) showed nearly 2 month improvement in progression free survival with addition of veliparib (120 mg BID on days -2 to 5) to standard doses of carboplatin and paclitaxel (compared to placebo added to carboplatin and paclitaxel) in patients with HER2 negative, metastatic breast cancer and germline BRCA 1/2 mutation.^40^

A unique aspect of our study is the inclusion of functional imaging using ^18^FLT-PET scans at early time points to non-invasively assess reduction in proliferation rate and compared this with ^18^FDG-PET scans. Increased uptake of ^18^FLT is seen in cells that express high levels of thymidine kinase 1, the key enzyme in the pyrimidine salvage pathway of DNA synthesis,^41^ hence correlating with increased cell proliferation. The SUV_max_ measurements on ^18^FLT-PET-CT has been shown to correlate with response to therapy in breast cancer^20^ and there is growing interest for the potential of early response monitoring in breast cancer using PET-based imaging modalities^21,22^. We performed ^18^FLT-PET scans at 3 early time points, (baseline, cycle 1 day 7 or 14, cycle 1 day 21, and cycle 3 day 1) as a tool to determine the impact of dose and schedule of veliparib on the proliferation rate of metastatic sites and we found that that the SUV_max_ on ^18^FLT-PET scan did not vary with dose or schedule (7 vs 14 vs 21 days) of veliparib. We demonstrated that among responders, drop in ^8^FLT uptake is rapid in many patients – within 7 days – implicating a potential early imaging marker of response. Two other studies in metastatic and primary breast cancer have demonstrated that changes in FLT-uptake in the primary or metastatic sites after one dose chemotherapy correlated with late tumor responses.^42,43^ By performing ^18^FLT-PET imaging along with ^18^FDG-PET imaging, this rich dataset also allowed comparison of PET radiotracers – we found that while ^18^FLT-PET and ^18^FDG-PET were overall correlated, there are differences and we are currently evaluating whether these modalities may be complementary. Our study has demonstrated that performing serial ^18^FLT-PET scans is feasible without adverse effects and can be a non-invasive tool to assess early response and proliferation rates.

In conclusion, our phase I, dose finding study of varying dose and schedule of veliparib along with carboplatin identified 250 mg BID daily as the recommended phase II dose and demonstrated safety and tolerability in patients with sporadic and BRCA mutated TNBC and in patients with HR positive MBC who had a functional deficiency of FA pathway as detected by FATSI assay. Furthermore, our study showed that use of novel functional FLT-PET-imaging as a tool to assess reduction in proliferative rate in the tumor and early response is feasible. Single agent PARPi, olaparib and talazoparib are approved for the management of BRCA mutated MBC. Our study provides rationale to study platinum/PARPi combination in other tumor subtypes as well.

## Data Availability

All clinical trial data will be deposited in accordance with CTEP, NCI, and NIH guidelines.

## DATA AVAILABILITY

All clinical trial data will be deposited in accordance with CTEP, NCI, and NIH guidelines.

## ACKNOWLEDGEMENTS

The study is supported by U01 CA076576. RW is supported by Translational Grant K12 CA133250 in experimental therapeutics from the National Cancer Institute and Cancer Clinical Investigator Team Leadership Award P30CA016058-42S2. DGS is supported by Susan G. Komen Career Catalyst Research Award.

## AUTHOR CONTRIBUTIONS

All authors made substantial contributions to the intellectual content of this work via conception, design, data acquisition, analysis, and/or interpretation, in addition to drafting and/or critical revision of the manuscript. All authors provided their final approval of the manuscript.

